# Development of Interactive Nomograms for Predicting Short-Term Survival in ICU Patients with Aplastic Anemia

**DOI:** 10.1101/2025.02.25.25322896

**Authors:** Junyi Fan, Shuheng Chen, Elham Pishgar, Greg Placencia, Maryam Pishgar

## Abstract

**Background:** Aplastic anemia is a severe hematologic disorder marked by pancytopenia and bone marrow failure. ICU admission often reflects disease progression or complications requiring critical care. Predicting short-term survival in these patients is vital for individualized treatment and resource optimization. Nomograms provide a practical tool for integrating clinical parameters, offering accurate visualized survival predictions to guide decision-making in patients with aplastic anemia in the ICU.

**Methods:** Using the MIMIC-IV database, we identified ICU patients diagnosed with aplastic anemia. From thousands of available variables, we extracted data across five dimensions: demographic, synthetic indicators, laboratory events, comorbidities, and drug usage. Based on existing studies of aplastic anemia, more than 400 variables were further refined and machine learning techniques were applied to identify the seven most effective predictors for modeling. Preprocessing was performed using machine learning approaches, and the feasibility of these predictors was validated through additional classification and regression models, the verification method is AUROC. Furthermore, external validation was performed using data from the eICU Collaborative Research Database to assess the generalizability of our models.The interactive nomograms were constructed using logistic regression (LR) to predict mortality rates at 7 days, 14 days, and 28 days in patients with aplastic anemia.

**Results:** A total of 1,662 patients diagnosed with aplastic anemia were included in this study, with a 7:3 ratio split into training and testing cohorts. The logistic regression model demonstrated strong predictive performance, achieving AUC values of 0.8227, 0.8311, and 0.8298 for 7-day, 14-day, and 28-day mortality predictions, respectively. External validation using the eICU database further confirmed the model’s generalizability, with AUC values of 0.7391, 0.7119, and 0.7093. These results highlight the model’s stability and effectiveness in predicting short-term survival in aplastic anemia patients.

**Conclusion:** A set of seven predictors, led by APS III, proved effective for modeling short-term survival in aplastic anemia patients. Using these predictors, Cox and logistic regression models generated nomograms that accurately predict 7-day, 14-day, and 28- day mortality. These tools can support clinicians in personalized risk assessment and decision-making.

## Introduction

Aplastic anemia is a rare but life-threatening hematological disorder characterized by bone marrow failure, leading to reduced hematopoietic stem cells and pancytopenia [1]. In Western countries, the incidence of aplastic anemia is approximately 2–3 cases per million people annually, while in East Asian countries, the incidence is higher, reaching 5–7 cases per million [2]. Without timely treatment, the disease often results in high mortality due to complications such as infections, bleeding, or organ failure [3]. Aplastic anemia severely impacts patients’ quality of life and poses a significant burden on healthcare systems [4].

The core pathological mechanism of aplastic anemia involves the depletion and dysfunction of hematopoietic stem cells, primarily driven by immune-mediated processes. Studies have shown that cytotoxic T lymphocytes secrete interferon-*γ* (IFN-*γ*) and tumor necrosis factor-*α* (TNF-*α*), which inhibit the proliferation and differentiation of hematopoietic stem cells [5]. In addition, genetic predisposition and environmental factors, such as exposure to certain drugs, chemicals, and viral infections, can further contribute to disruption of the bone marrow microenvironment and exacerbate hematopoietic dysfunction [2].

Aplastic anemia presents significant diagnostic challenges, particularly in its early stages, as it can mimic other conditions causing pancytopenia, such as myelodysplastic syndromes or malignant hematological diseases. Bone marrow biopsy remains the gold standard for diagnosis, but is invasive and often uncomfortable for patients [5]. In addition, the diverse clinical presentations of aplastic anemia, ranging from mild anemia to severe bleeding tendencies, often lead to misdiagnosis or delayed diagnosis. For example, studies have reported that approximately 10% of patients initially misdiagnosed with immune thrombocytopenia were later confirmed to have aplastic anemia [3]. These challenges underscore the need for improved diagnostic tools to enable early and more accurate diagnosis of the disease.

Treatment of aplastic anemia consists mainly of immunosuppressive therapy (IST) and hematopoietic stem cell transplantation (HSCT). For non-severe cases or patients ineligible for transplantation, IST based on anti-thymocyte globulin (ATG) and cyclosporine is the first-line treatment, achieving an overall response rate of 60–80% [6]. However, around 30% of patients may experience relapse or refractory disease [7]. For severe cases eligible for HSCT, it remains the only potentially curative option, with a five-year survival rate exceeding 80% when a matched donor is available [6]. Accurate prediction of short-term survival in aplastic anemia patients in the ICU can further enhance treatment outcomes by allowing personalized therapeutic strategies, guiding the timing and selection of interventions, and optimizing the use of critical care resources. In recent years, the introduction of hematopoietic stimulating agents, such as eltrombopag, and immune modulators, has improved outcomes for patients with aplastic anemia, although effective strategies for refractory cases remain an ongoing area of research [8].

Recent advances in machine learning have shown promise in improving the diagnosis and management of aplastic anemia. For example, Wang et al. developed a convolutional neural network model capable of automatically distinguishing between aplastic anemia, myelodysplastic syndromes, and acute myeloid leukemia based on bone marrow smear images, achieving high precision and potentially reducing diagnostic delays [9]. In addition, machine learning models have been explored for pre-transplant risk stratification in patients with severe aplastic anemia, which helps improve treatment planning and improve patient outcomes [10]. These developments suggest that the integration of machine learning approaches can provide clinicians with valuable tools for more accurate diagnosis and personalized treatment strategies in aplastic anemia.

## Methodology

### Data Source

The Medical Information Mart for Intensive Care IV (MIMIC-IV) is a comprehensive and freely accessible database that compiles deidentified health data from patients admitted to intensive care units (ICU) at Beth Israel Deaconess Medical Center between 2008 and 2019. This resource covers a wide range of information, including demographics of the patient, vital signs, laboratory results, medications, and clinical notes. The primary purpose of MIMIC-IV is to facilitate critical care research and support the development of predictive models by providing a rich data set that reflects real-world clinical scenarios [11].

By integrating data from these comprehensive datasets, we can capture detailed clinical measurements and observations across a heterogeneous patient population. This diversity ensures that the data are representative of various demographics and conditions of the patients, thereby improving the generalizability of the research findings. For studies focusing on rare diseases such as aplastic anemia, MIMIC-IV offers valuable information by providing access to pertinent patient records, which can be instrumental in understanding disease patterns, treatment responses, and outcomes [12].

For external validation, we apply e-ICU. The Collaborative Research Database eICU is a multicenter critical care database comprising deidentified health data associated with more than 200,000 ICU admissions in the United States [13].

### Study Population

The population selection focuses on ICU patients diagnosed with aplastic anemia or related conditions using ICD-9 codes (**284**) and ICD-10 codes (**D60, D61**). Patients are filtered to include only those with an ICU stay of at least one day and who survived beyond the first 24 hours of ICU admission. Additionally, only patients aged between 18 and 90 years are included. For patients with multiple stays in the ICU, only the first admission is considered, ensuring one record per subject. This approach ensures a clinically meaningful cohort by excluding short-term stays, early deaths, and patients outside the target age range. The final data set includes subject identifications, hospital admissions, stays in the ICU and timestamps of admission, discharge, and death for further analysis. The extraction workflow for the study population is shown in Fig 1

**Fig 1.**
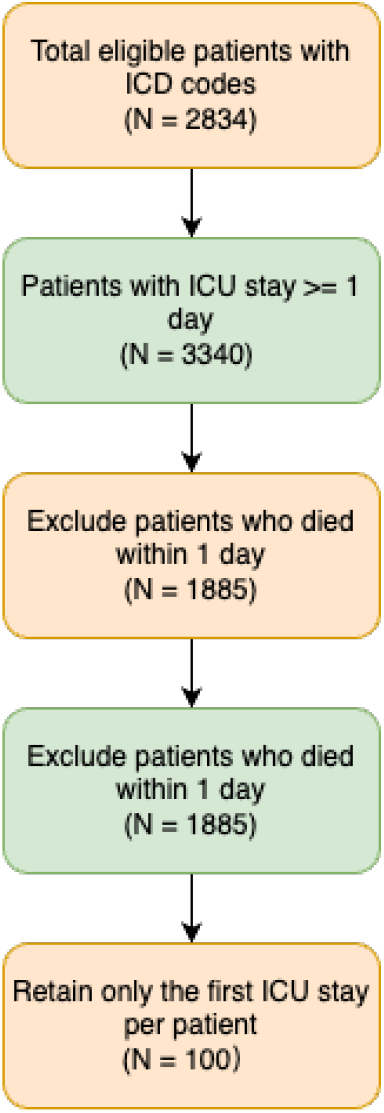
Criterion of study population extraction

### Feature Selection

In this study, the selection of characteristics was carried out in two sequential steps to identify the most relevant predictors to model short-term survival in ICU patients with aplastic anemia. Initially, a filter-based method, SelectKBest, was applied to narrow down the pool of over 400 candidate variables to the top 50 features based on their statistical association with the outcome variable.

SelectKBest ranks features based on a chosen statistical metric, such as the F-test for classification tasks:

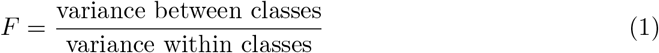

where a higher *F* value indicates stronger relevance to the target variable. Features with the top *K* scores are retained, effectively removing noise and redundant information. By applying SelectKBest, we ensured that only the most statistically significant features contributed to downstream modeling. This step streamlined our predictive framework, enabling better generalization without sacrificing essential clinical information. The reduced feature set facilitated more efficient training and improved model interpretability while preserving predictive performance [14].

Subsequently, a wrapper-based method, Recursive Feature Elimination (RFE), was employed to further refine the feature set. Recursive Feature Elimination (RFE) is an iterative feature selection technique that eliminates the least important features based on model performance, optimizing predictive power while reducing dimensionality. At each iteration, a model assigns weights *β*_*i*_ to features, and the least significant feature is removed:

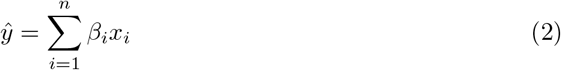

where *β*_*i*_ represents the feature importance. This approach iteratively evaluates subsets of features by training and testing the model performance, ultimately identifying the optimal set of 7 features that contribute the most to predictive accuracy [15].

At the same time, we take expert advice on the selecting process. The final selected features were: *APS III, Anion Gap, Base Excess, Bicarbonate, Fibrinogen, Functional, Lactate Dehydrogenase (LD)*, and *platelet count*. These variables reflect key clinical and biochemical parameters associated with patient outcomes and serve as the foundation for building the predictive nomogram [16].

The selected features are shown in Table 1

**Table 1.**
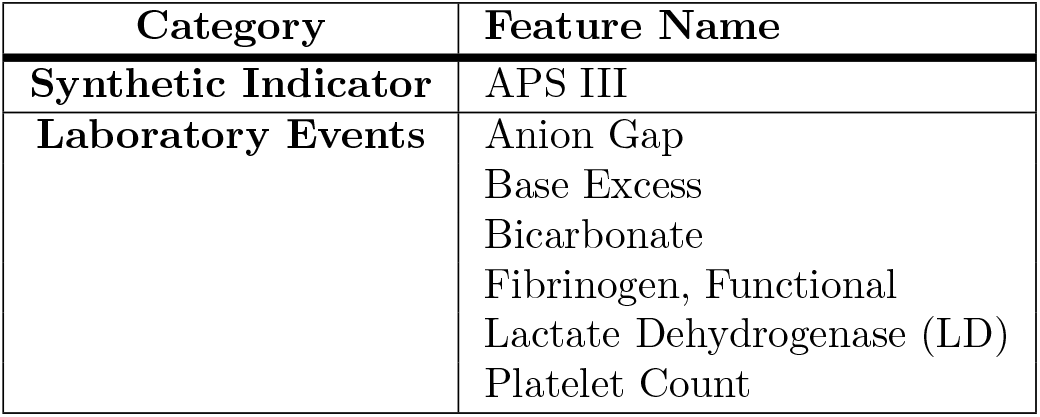
Selected clinical features.

Among these, APS III is used to gauge the severity of bone marrow failure, providing an early prognostic indicator [17]. The APS III score is calculated using 17 physiological variables, each weighted according to its deviation from the normal range. The final score ranges from 0 to 252, where a higher score indicates greater severity of the illness.

Mathematically, APS III is expressed as:

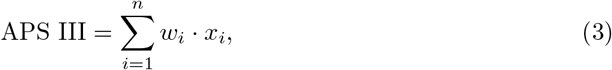

where *x*_*i*_ represents the observed value of the *i*^*th*^ physiological variable, and *w*_*i*_ is the corresponding weight assigned based on the deviation from normal values. The physiological parameters include heart rate, mean arterial pressure, respiratory rate, oxygenation levels (PaO_2_/FiO_2_), blood pH, sodium, potassium, glucose, creatinine, blood urea nitrogen, white blood cell count, hematocrit, temperature, urine output and Glasgow Coma Scale (GCS).

The anions gap has also been associated with metabolic stress in aplastic anemia, where elevated levels may reflect underlying complications [18].

Base excess, which serves as a marker of acid-base balance, has been reported to correlate with disease progression in marrow suppression [19], while bicarbonate levels can offer information on body compensatory responses during bone marrow insufficiency [20].

Here is the calculation of base excess:

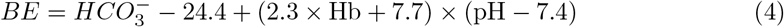

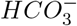 : Bicarbonate concentration in blood, measured in milliliters per liter (mEq / L).

- 24.4 : Normal bicarbonate concentration at standard conditions (mEq/L).
- Hb : Hemoglobin concentration in the blood, measured in grams per deciliter (g/dL).
- pH : Arterial blood pH, which represents the acid-base status of the patient.
- 2.3 : Coefficient related to the buffering capacity of hemoglobin.
- 7.7 : Constant derived from standard physiological conditions.
- 7.4 : Normal arterial blood pH.

Fibrinogen, a key clotting factor, is often monitored to assess the risk of coagulopathy, which can be exacerbated by reduced platelet production [21].

Furthermore, lactate dehydrogenase (LD) frequently serves as a marker of tissue turnover, with elevated values suggesting more aggressive disease activity [22].

Finally, Platelet Count remains a central measure for evaluating both disease severity and treatment response, where chronic thrombocytopenia signals poorer clinical outcomes [23].

The variance inflation factor (VIF) is a statistical measure that is used to detect multicollinearity among independent variables in a regression model. Quantifies how much the variance of a regression coefficient is inflated due to collinearity with other predictors. A VIF value greater than 5 is often considered indicative of high multicollinearity, which can compromise the reliability of the model by inflating standard errors and reducing interpretability [24].

In this study, all selected predictors demonstrated VIF values Fig 2 well below the commonly accepted threshold of 5, indicating minimal multicollinearity between variables. This confirms that the selected features, including *APS III, Anion Gap, Base Excess, Bicarbonate, Fibrinogen, Functional, Lactate Dehydrogenase (LD)*, and *platelet count*, are independent and suitable for use in predictive modeling without significant redundancy.

**Fig 2.**
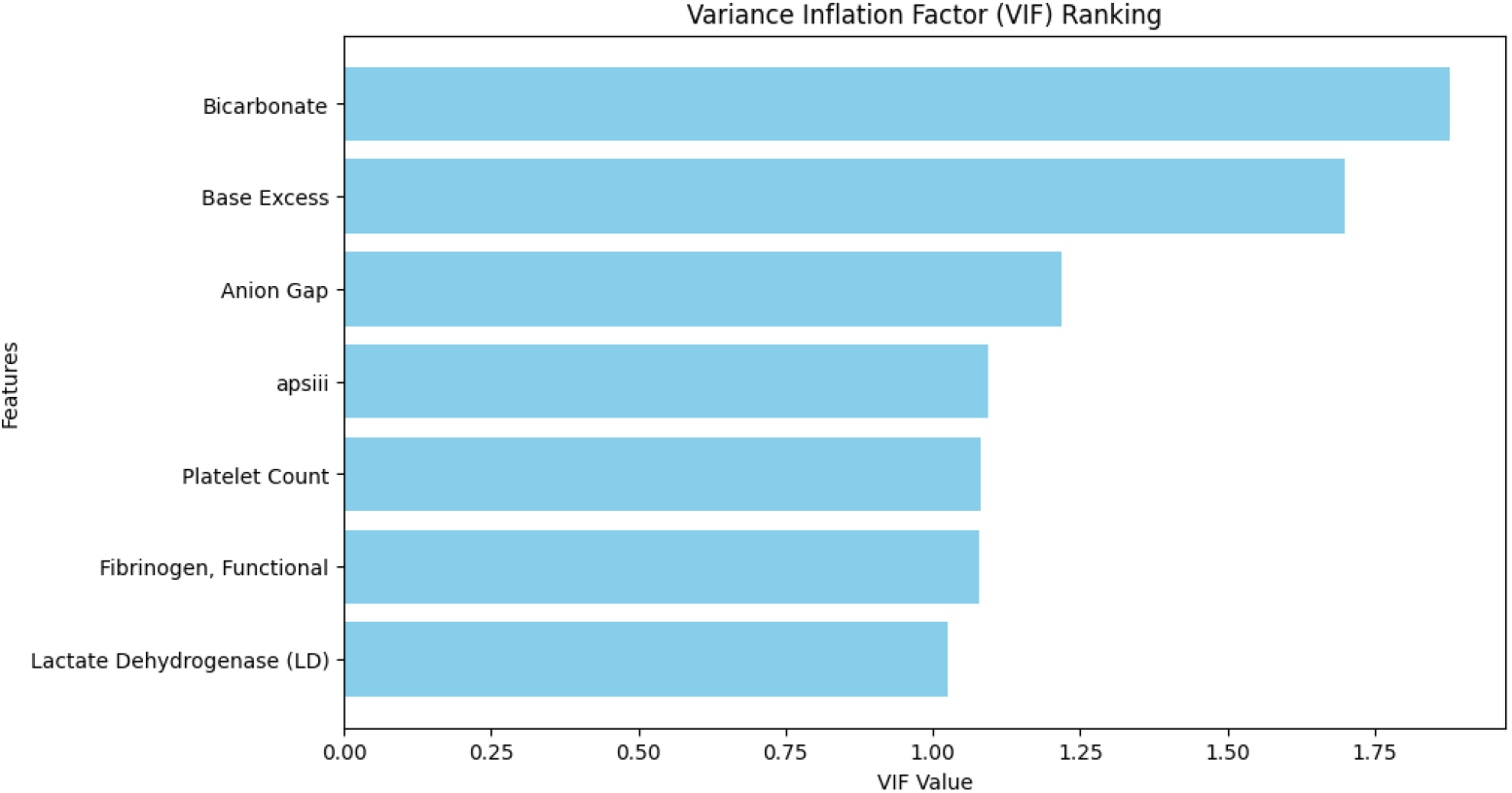
Variance Inflation Factor of selected features.

### Custom SMOTE for imbalance data

To address the imbalance in continuous target variables, we propose a **custom SMOTE (Synthetic Minority Oversampling Technique) for continuous data**, which enhances traditional SMOTE by incorporating weighted sampling based on predefined thresholds.

#### Algorithm Overview

##### 1. Weight Assignment

The continuous target variable *Y* is segmented into intervals based on user-defined thresholds {*T*_1_, *T*_2_, …, *T*_*n*_}, with corresponding weights {*w*_1_, *w*_2_, …, *w*_*n*+1_}. Each sample is assigned a weight on the basis of the interval in which it falls.

##### 2. Weighted Sampling

The probabilities for resampling are computed as:

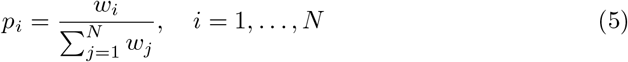

where *N* is the number of original samples.

##### 3. Synthetic Sample Generation

Additional samples are generated through interpolation. For each selected sample *y*_*i*_, a set of *k* randomly chosen nearest neighbors is used to calculate an interpolated value:

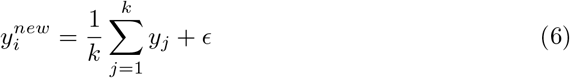

where *ϵ* ∼ *U* (−*δ, δ*) represents a small uniform noise to introduce variability.

##### 4. Data Augmentation

The newly synthesized samples are appended to the original dataset, yielding an expanded dataset (*X*_*resampled*_, *Y*_*resampled*_).

### Experimental Parameters

In our experiment, we set thresholds as {7, 35} with weights {40, 20, 1} and generated 2000 additional samples.

## Results

Table 2 presents the size of the data set before and after applying our method.

**Table 2.** Dataset dimensions before and after applying Custom SMOTE.

| Dataset | Original Shape | Resampled Shape |
| --- | --- | --- |
| $X$ | $(m, d)$ | $(m + 2000, d)$ |
| $Y$ | $(m, )$ | $(m + 2000, )$ |

**Table 3.** Comparison of population features between training and test datasets.

| Variable | Unit | Training Mean (SD) | Test Mean (SD) | P-value |
| --- | --- | --- | --- | --- |
| APS III | - | 51.8 (18.5) | 52.9 (18.8) | 0.273 |
| Anion Gap | mEq/L | 12.9 (2.6) | 12.9 (2.6) | 0.913 |
| Base Excess | mEq/L | -1.1 (3.5) | -1.2 (3.4) | 0.712 |
| Bicarbonate | mEq/L | 23.8 (3.2) | 23.6 (3.3) | 0.311 |
| Fibrinogen | mg/dL | 337.4 (143.9) | 345.2 (147.8) | 0.324 |
| LD | U/L | 283.1 (112.9) | 291.6 (119.1) | 0.177 |
| Platelet Count | 10<sup>9</sup>/L | 95.8 (54.4) | 89.9 (51.5) | 0.037 |

Values are presented as mean (standard deviation). APS III: Acute Physiology Score III; LD: Lactate Dehydrogenase.

This method effectively balances the distribution of continuous targets while maintaining statistical coherence, reducing bias towards underrepresented regions in the dataset.

### Modeling

In this study, the main objective was to develop robust predictive models for short-term survival in ICU patients with aplastic anemia. Two main models were applied to construct interpretable *nomograms*: Logistic Regression (LR) Logistic Regression is a widely used statistical model for binary classification tasks, such as predicting survival (yes/no) within a specific time period. Estimates the probability of an event occurring based on linear combinations of independent variables. The simplicity and interpretability of LR make it a favored choice for constructing nomograms, where the model coefficients are used to assign scores for individual risk assessment.

Logistic regression probability estimation model:

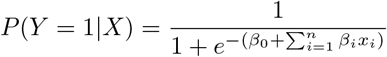

To predict 7-day, 14-day, and 28-day mortality of ICU patients with aplastic anemia, we use the following models.

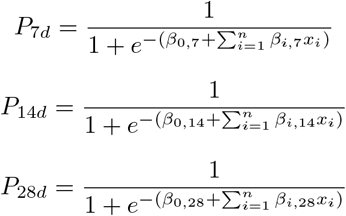

where *x*_*i*_ represents the input features:

Cox regression, on the other hand, is a semiparametric model commonly used in survival analysis. Estimates the hazard ratio for a given set of covariates without making strong assumptions about the underlying hazard function. This model allows for the prediction of event probabilities over time, making it highly suitable for creating time-dependent nomograms. In this study, Cox regression was used to predict survival probabilities of 7 days, 14 days, and 28 days.

To ensure the robustness and feasibility of the selected features, as well as to benchmark against traditional models, several machine learning algorithms were employed. These include Random Forest (RF), XGBoost (Extreme Gradient Boosting), LightGBM, Neural Networks (NN) and AdaBoost. These machine learning models served primarily to validate the findings from LR and Cox regression, ensuring that the selected features and predictive models were both interpretable and highly predictive.

## Results

### Statistical Comparison

A statistical comparison between the training and test data sets was made for key clinical variables. Table **??** presents the mean and standard deviation (SD) for each variable in both datasets, together with the corresponding p-values of the independent two-sample t-tests.

APS III did not show significant differences between the training (mean = 51.8, SD = 18.5) and the test sets (mean = 52.9, SD = 18.8, p = 0.273). Similarly, metabolic parameters such as adenosine equilibria, excess base, and bicarbonate levels did not show statistically significant differences (p-values = 0.913, 0.712, and 0.311, respectively).

Coagulation-related variables, including fibrinogen (p = 0.324) and Platelet Count (p = 0.037), were analyzed. Although fibrinogen levels remained comparable between the groups, platelet count exhibited a marginally significant difference (mean = 95.8 vs 89.9, p = 0.037). Lactate dehydrogenase (LD), a biomarker of cellular injury, also did not show significant variation (p = 0.177).

In general, the distribution of clinical variables suggests a high degree of similarity between the training and test sets, ensuring a consistent evaluation framework for model performance.

### Model Performance

The Logistic Regression (LR) model demonstrated consistent predictive performance at all time points. For the prediction of survival at 7 days, the model achieved an AUROC of 0.823 (95% CI: 0.737–0.909). Similarly, for 14-day and 28-day survival predictions, the AUROC values were 0.831 (95% CI: 0.766–0.896) and 0.830 (95% CI: 0.773–0.887), respectively, indicating strong discriminatory power over time. Although Precision-Recall AUC values (PR AUC) increased from 0.323 at 7 days to 0.516 at 28 days, the model consistently provided reliable predictions for short-term survival. The AUROC curves for our proposed model are shown in Fig 3

**Fig 3.**
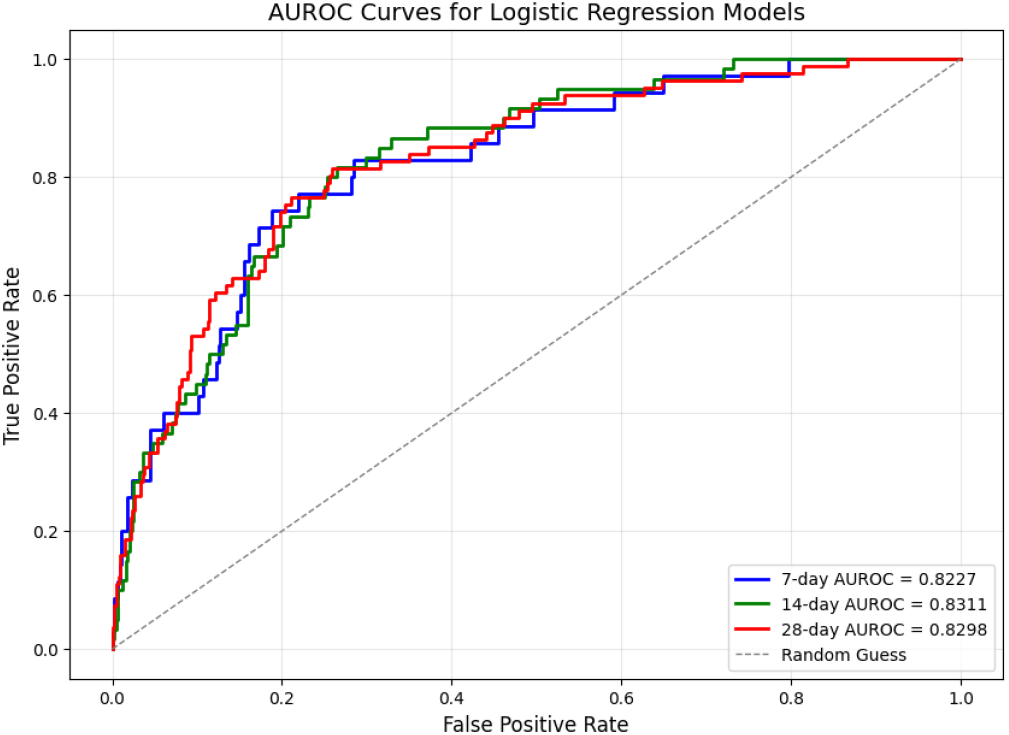
AUROC-curves for test set of LR Models

Although several machine learning models, including Random Forest (RF), Neural Networks (NN), LightGBM, and XGBoost, were evaluated to predict mortality at time points of 7 days, 14 days, and 28 days, their overall performance was found to be inferior to the Logistic Regression (LR) model. Despite achieving high specificity, these models consistently demonstrated zero sensitivity at all time points, reflecting the challenges of correctly predicting positive cases. LR outperformed these models in both AUROC and sensitivity, highlighting its robustness and suitability to develop reliable nomograms for the prediction of short-term mortality in ICU patients with aplastic anemia. Other models are shown in Fig 4

**Fig 4.**
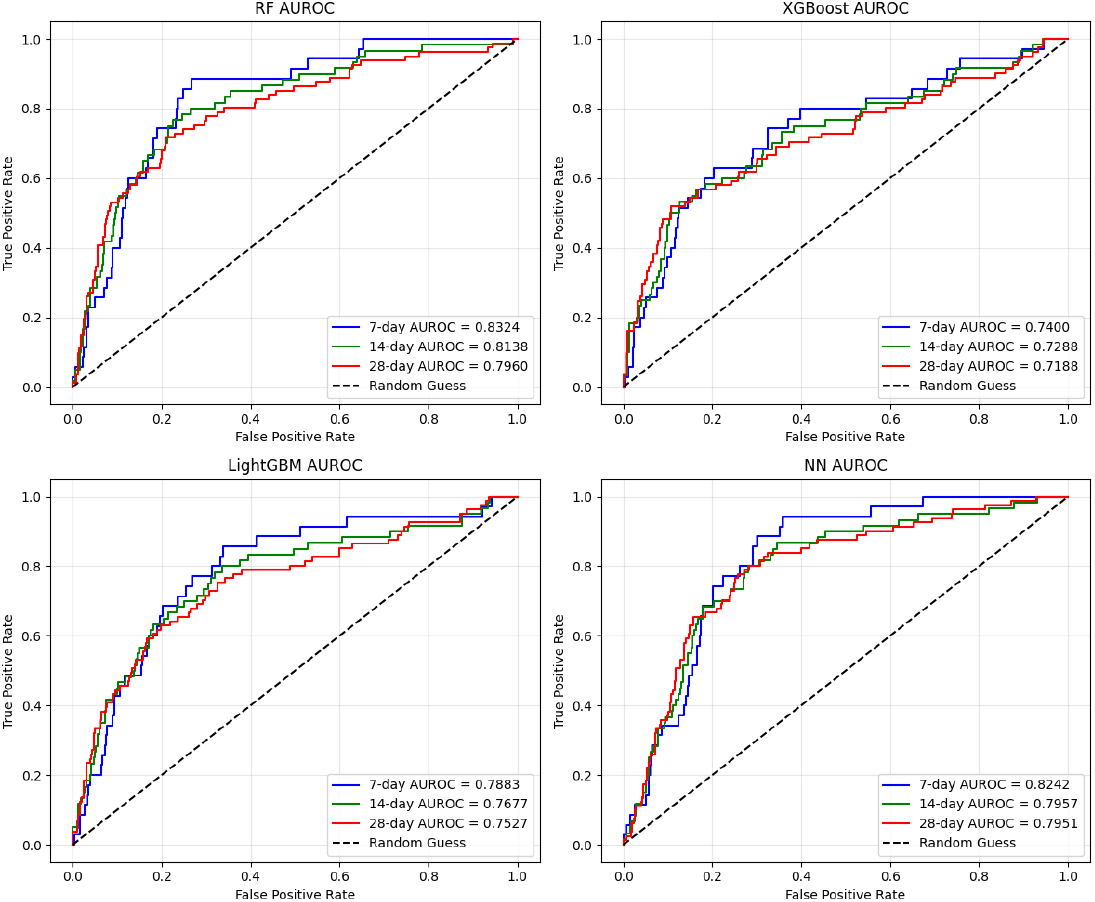
AUROC-curves for test set of baseline Models

**Fig 5.**
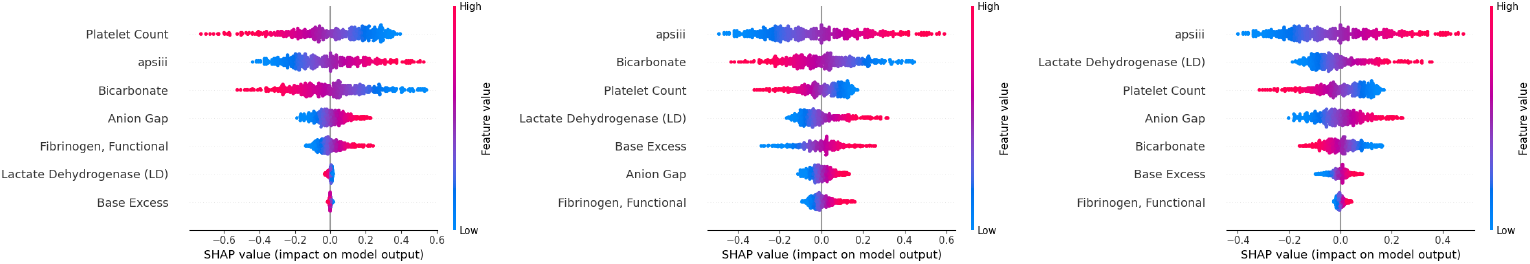
SHAP Summary Plots for Different Mortality Thresholds

**Fig 6.**
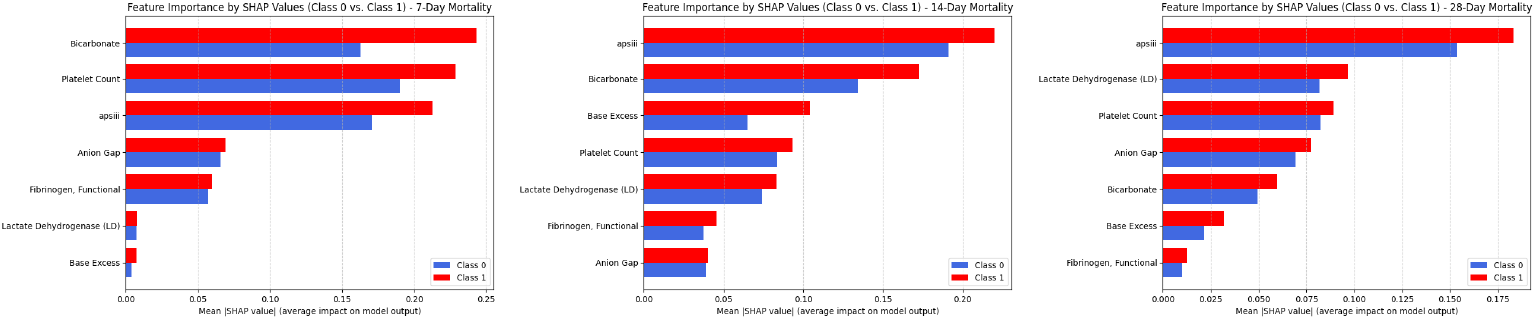
SHAP Summary Plots for Different Mortality Thresholds

Our Cox regression model significantly outperformed the reference study by Tu et al. [25], achieving a C index of 0.761 compared to 0.643, an improvement of 18. 3%. This enhancement is attributed to our refined feature selection, incorporating machine learning techniques, and advanced data preprocessing, such as KNN imputation and outlier handling. These methodologies ensured a robust data set and a model capable of delivering more accurate predictions, demonstrating its clinical utility to guide ICU management for patients with aplastic anemia.

### SHAP Analysis

SHAP (SHapley Additive exPlanations) is a game-theoretic approach to interpreting machine learning models. It assigns each characteristic an importance score based on its contribution to the prediction of the model, ensuring a consistent and fair attribution of predictive power. SHAP values help us to understand not only which features influence predictions, but also the direction of their effect. In this study, SHAP analysis was used to examine the impact of various clinical variables on 7-day, 14-day, and 28-day mortality predictions using logistic regression models.

Across the three models, APS III, bicarbonate, and platelet count consistently emerged as key predictors. APS III, a widely used ICU severity score, was of high importance, while bicarbonate levels reflected metabolic disturbances linked to poor outcomes. Platelet count was crucial in patients with aplastic anemia, as thrombocytopenia increases the risk of bleeding and mortality.

The importance of features varied between models. In the 7-day model, bicarbonate and platelet count dominated, highlighting acute metabolic and hematological effects on short-term survival. In the 14-day and 28-day models, APS III became more influential, indicating systemic severity’s growing role. The lactate dehydrogenase and anion gap also fluctuated, reflecting evolving metabolic and inflammatory responses.

These results, which are shown in the study, suggest that while some prognostic factors remain critical, their relative impact changes over time. Continuous monitoring and dynamic risk assessment are essential for patients in the ICU of aplastic anemia. SHAP analysis improves interpretability, supporting data-driven clinical decisions and personalized treatment strategies.

### Permutation Importance Analysis

Permutation importance is a widely used technique to assess the contribution of individual features in predictive models. The method involves randomly shuffling the values of a single feature while keeping others unchanged, measuring the corresponding decline in model performance. The importance of feature *X*_*j*_ is computed by comparing the baseline AUROC *AUC*_base_ with the AUROC after shuffling *AUC*_perm_:

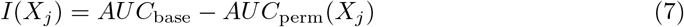

where a larger *I*(*X*_*j*_) indicates that the feature has a stronger influence on the predictive performance of the model. This process is repeated multiple times to obtain a robust estimate of the importance of the feature.

The permutation importance analysis, shown in Fig 7 revealed that APS III was the most influential predictor in all three models, highlighting its role as a key severity indicator in ICU patients with aplastic anemia. Platelet count and bicarbonate consistently ranked among the top predictors, highlighting the impact of hematologic status and metabolic balance on short-term survival. Anion gap and lactate dehydrogenase (LD) showed moderate importance, suggesting their relevance in systemic metabolic and inflammatory responses, particularly at later time points.

**Fig 7.**
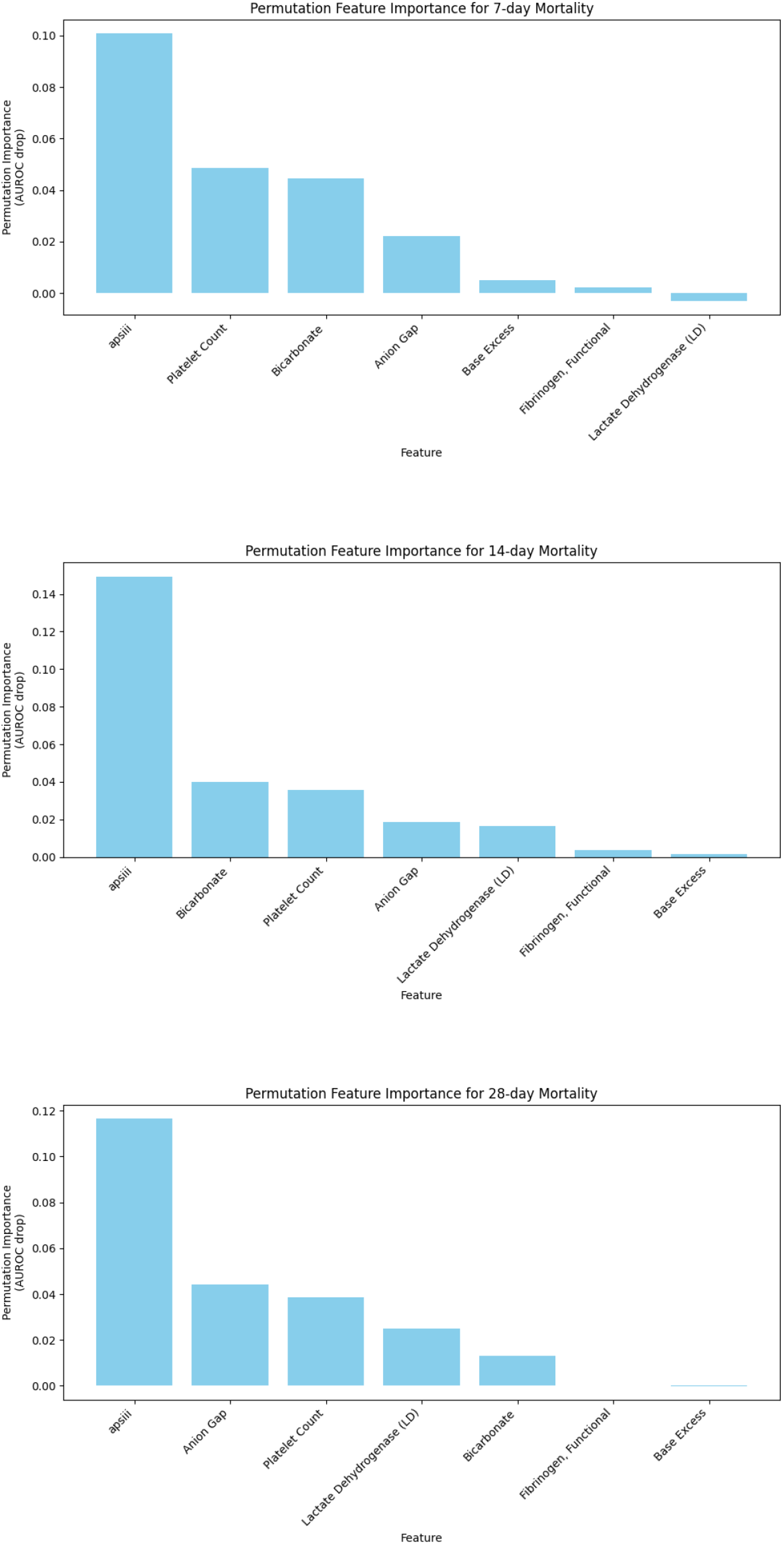
Permutation Importance of Features for 7-day, 14-day, and 28-day Mortality Prediction

Features such as base excess and fibrinogen had minimal impact, indicating their limited predictive value in this cohort. While APS III remained dominant across all models, the relative importance of metabolic indicators (bicarbonate, anion gap) increased over time, possibly reflecting a shift from acute physiological instability to longer-term metabolic deterioration in ICU patients.

### External Validation

External validation, shown in Fig 8 is a critical step in assessing the generalizability of predictive models. To validate our logistic regression (LR) model externally, we applied the same data extraction methodology to the eICU Collaborative Research Database and identified 452 ICU patients diagnosed with aplastic anemia. This sample size closely matches that of the test dataset, ensuring a comparable evaluation. The same seven key predictors were extracted, and the pre-trained LR model was applied to assess its performance in an independent cohort. The model demonstrated robust predictive capability, achieving AUC values of 0.7391, 0.7119, and 0.7093 for 7-day, 14-day, and 28-day mortality predictions, respectively. These results indicate that despite the differences in patient demographics, institutional protocols, and clinical practices between MIMIC-IV and the eICU, the model maintains good discriminatory power.

**Fig 8.**
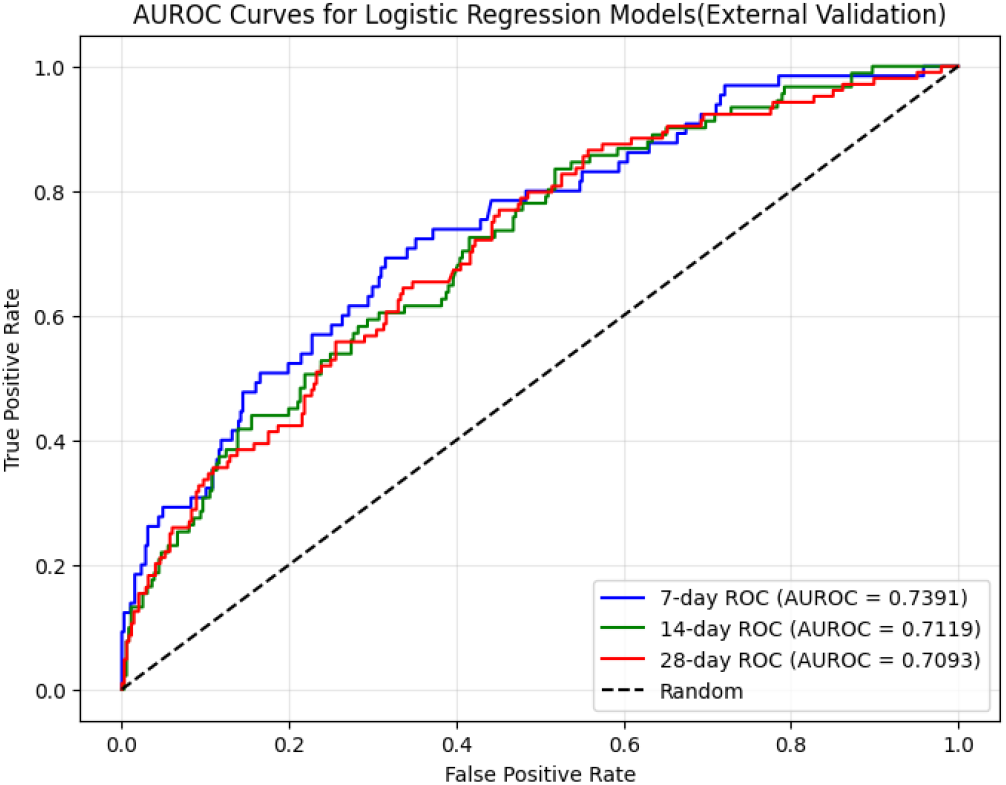
AUROC-curves for external validation data set of LR Models

The findings further support the reliability of the model for real-world applications and highlight the potential of machine learning-driven predictive tools to aid clinical decision making for ICU patients with aplastic anemia.

### Nomograms

The nomograms, shown in Fig 9, Fig 10 and Fig 11 illustrate the predicted short-term survival rates of ICU patients with aplastic anemia for 7, 14, and 28 days, integrating key clinical variables such as APS III, anions gap, base excess, bicarbonate, fibrinogen, lactate dehydrogenase, and platelet count. These tools allow clinicians to estimate the risk of mortality by mapping patient-specific values onto a scoring scale. Across the three time frames, the total points are directly proportional to the probability of mortality, and APS III and Bicarbonate consistently contribute significantly to risk estimation. The 7-day nomogram focuses on acute survival predictions, showing relatively lower total score ranges, indicative of lower cumulative risk in the short term. In contrast, the 28-day nomogram reflects a broader score range, highlighting the accumulation of risk over time, with variables such as LD and fibrinogen exerting greater influence in longer-term survival predictions. In particular, while all three nomograms share a common structure, they differ in score calibration to reflect the impact of time on survival probabilities. These nomograms provide a practical framework for clinicians to stratify mortality risks and guide personalized interventions for AA patients in critical care, highlighting the importance of acute and long-term physiological factors in survival outcomes.

**Fig 9.**
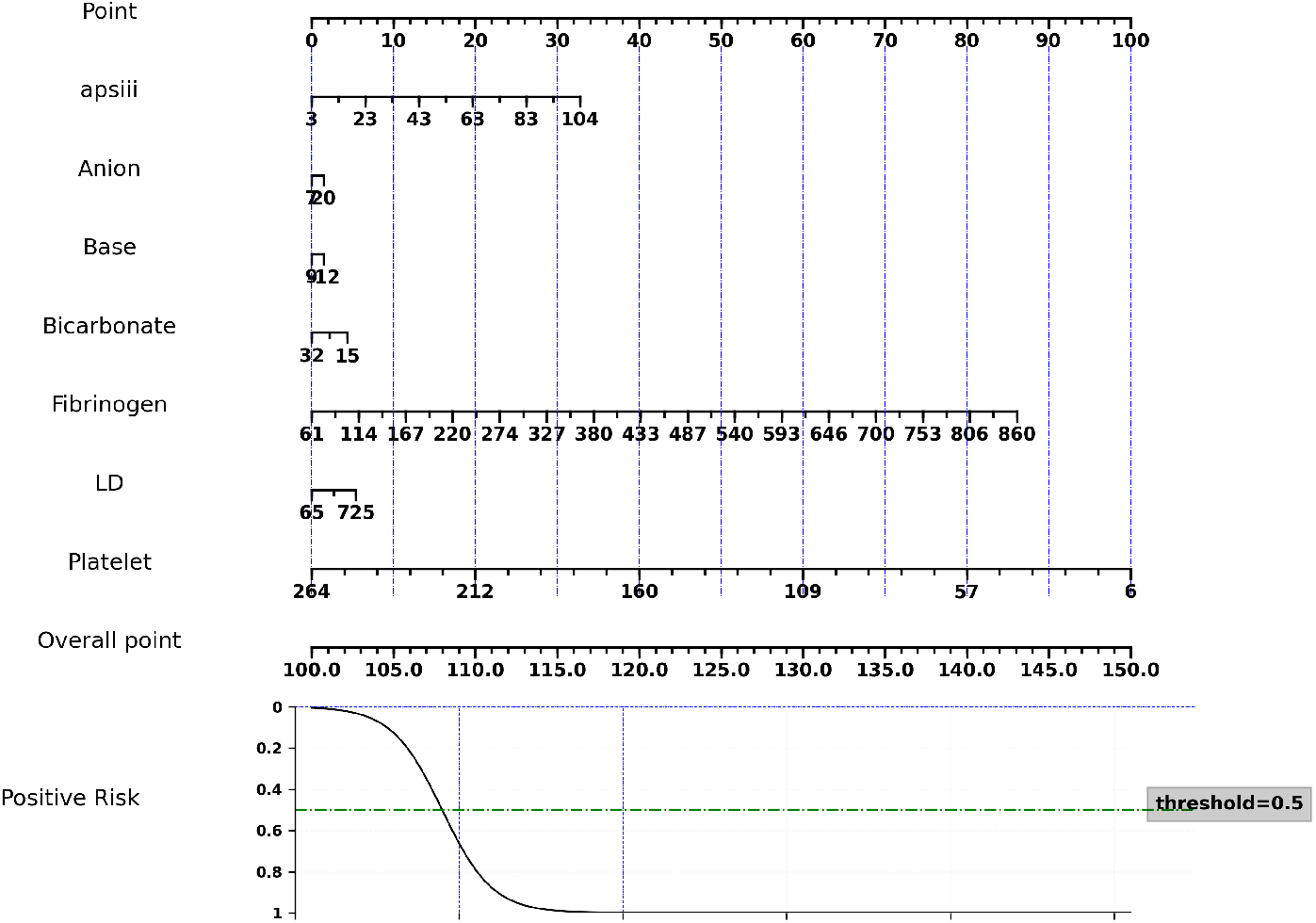
Nomogram of 7 days survival prediction

**Fig 10.**
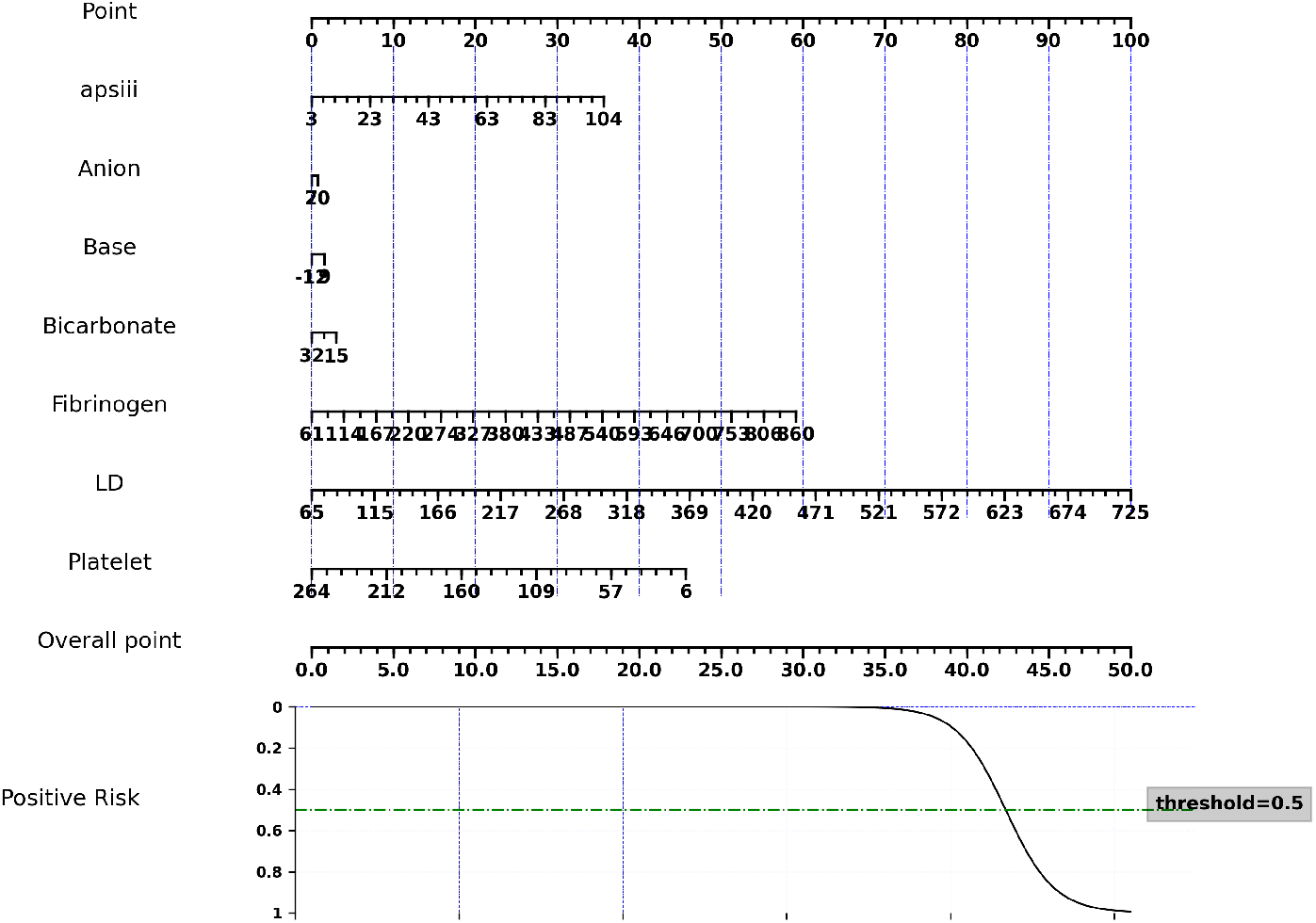
Nomogram of 14 days survival prediction

**Fig 11.**
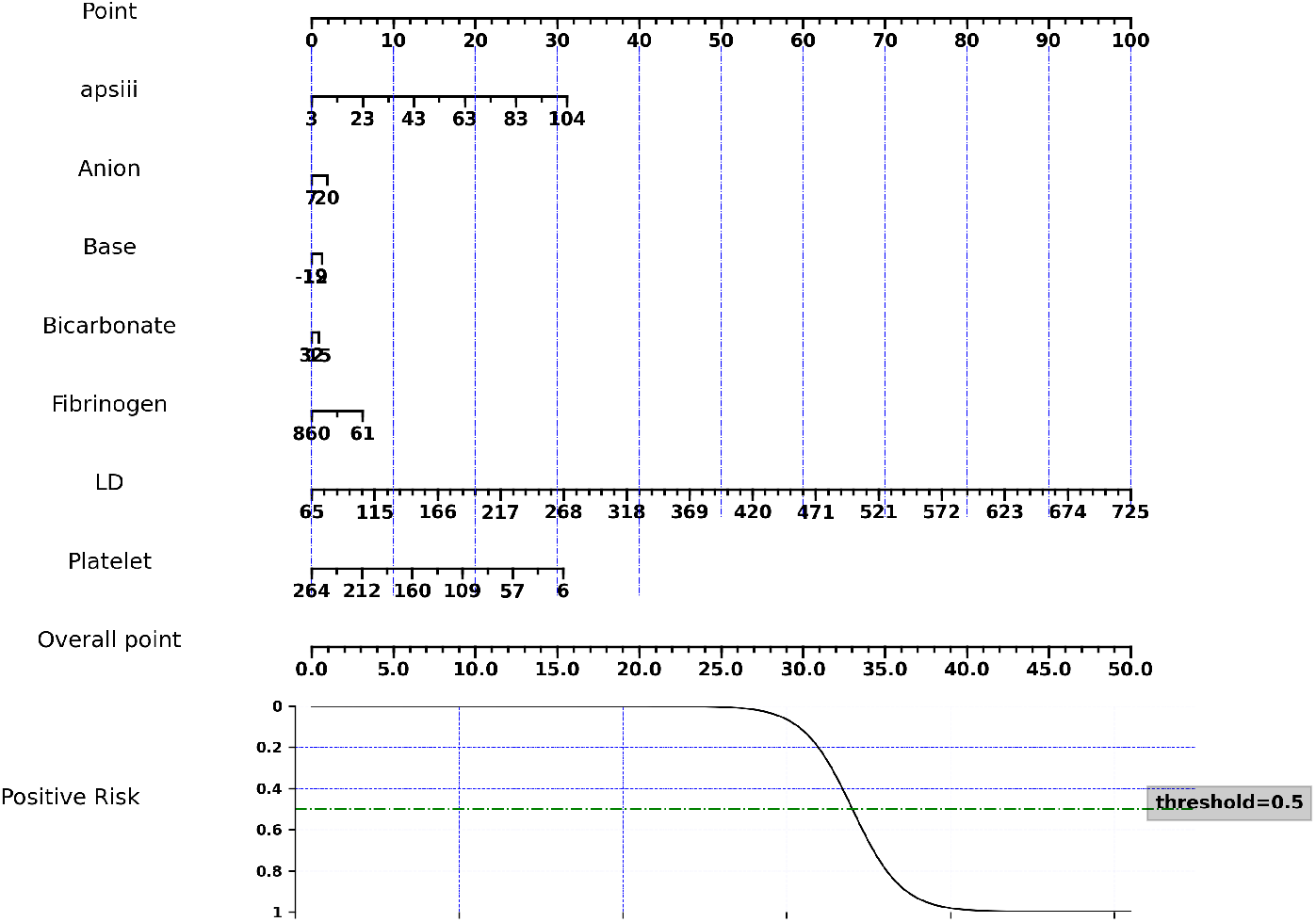
Nomogram of 28 days survival prediction

### Interactive Nomogram

An interactive nomogram was developed to predict ICU mortality for patients with aplastic anemia at 7, 14 and 28 days.

Patient data was extracted from the MIMIC-IV database for processing. Logistic regression was used to estimate the probability of mortality. An interactive web-based tool was developed using Dash and plotly to provide a user-friendly interface. A gauge-style nomogram was implemented to enable real-time risk assessment and visualization.

It is publicly available on GitHub at the following link: https://github.com/JunyiTim/Interactive-nomogram-for-ICU-patients-with-aplastic-anemia-

This tool provides a user-friendly interface for clinicians to estimate mortality risk based on patient-specific parameters, improving decision support in ICU settings.

## Discussion

### Model compliation Summary

Machine learning studies related to the diagnosis of aplastic anemia are currently limited. Compared to existing nomogram-based studies, our Cox regression model significantly outperformed the reference study by Tu et al. [25], achieving a C index of 0.761 compared to 0.643, reflecting an improvement of 18. 3% in predictive performance.

To enhance feature selection, we used RFECV (Recursive Feature Elimination with Cross-Validation) combined with clinical expertise, reducing the number of predictors from 50 to 7 key features. This approach used advanced machine learning techniques to identify valuable predictors while ensuring interpretability, making the model clinically meaningful.

To address class imbalance in the dataset, we developed a novel resampling method.

Custom SMOTE (Synthetic Minority Oversampling Technique) for continuous data.

Unlike traditional SMOTE, our approach incorporates weighted sampling based on predefined thresholds, generating sufficient positive cases in the training set, and improving model stability.

Although previous studies relied primarily on Cox regression, our study used multiple machine learning models, including logistic regression, AdaBoost, and tree-based algorithms, to validate performance. Each model consistently produced highly accurate predictions, demonstrating the robustness of our approach.

Finally, we performed external validation using the eICU database, confirming the generalizability of both our selected features and models. The results further support the applicability of our framework in predicting short-term mortality in ICU patients with aplastic anemia, reinforcing its potential clinical utility.

### Clinical Implications

Nomograms provide a rapid and interpretable method to estimate the survival probability of ICU patients, offering several advantages in medical diagnosis and clinical decision-making.

First, they enable individualized risk assessment by integrating multiple predictive variables into a visual representation, helping physicians make informed prognostic evaluations.

Second, nomograms enhance shared decision-making by allowing healthcare providers to communicate complex statistical predictions in an intuitive manner, facilitating discussions with patients and families regarding treatment options.

Third, they support triage and resource allocation, ensuring that high-risk patients receive timely interventions while optimizing ICU resource utilization.

Fourth, nomograms serve as a valuable research tool, allowing clinicians to compare different predictive models and validate their effectiveness in diverse populations.

Finally, these models contribute to personalized medicine, as their flexibility enables adaptation to specific diseases, improving precision in clinical predictions, and ultimately improving patient outcomes.

### Limitations and Future Directions

This study has several limitations that warrant further exploration. First, the models were developed using the MIMIC-IV single-center database, which may limit their generalizability to broader populations. External validation using the eICU database showed an approximate 10% decline in AUC performance compared to internal validation, indicating potential heterogeneity of the data set. Future research should incorporate data from multiple databases to allow cross-validation between diverse cohorts and improve the robustness of the models.

Second, while logistic regression and Cox regression provide interpretable results and facilitate the creation of nomograms, they remain relatively traditional. Despite their strengths in clinical applicability, these models can be affected by underlying multicollinearity issues, as analysis of variance inflation factor (VIF) alone cannot fully capture complex interdependencies between predictors. The presence of hidden correlations could impact model reliability, suggesting the need for additional feature engineering techniques.

Third, the diagnostic criteria and understanding of aplastic anemia continue to evolve. As medical research advances, the integration of novel biomarkers and emerging clinical indicators could further refine the predictive accuracy. In addition, most of the predictors used in this study are static values or represent averaged values over a specific time frame, which may not fully capture dynamic physiological changes. The lack of continuous monitoring data limits the ability to assess time-dependent fluctuations that could significantly impact survival predictions.

Future studies should explore the potential of advanced machine learning approaches, such as deep learning and time series models, to capture complex temporal patterns in ICU patients with aplastic anemia. Incorporation of recurrent neural networks (RNNs) or attention-based models could improve predictive performance by taking into account longitudinal variations in physiological parameters.

In addition, more sophisticated feature selection techniques could be applied to mitigate multicollinearity risks and enhance the interpretability of prediction models. Methods such as elastic net regularization and Bayesian variable selection may provide better feature stability while maintaining model accuracy.

Finally, expanding external validation efforts across diverse clinical datasets will be crucial to ensuring model robustness. Multicenter collaborations and federated learning frameworks may allow secure data sharing while addressing concerns about data set bias. By incorporating a larger patient population and accounting for regional treatment variations, future research can establish more generalizable predictive tools to support ICU decision making.

## Conclusion

This study developed predictive models for short-term survival in ICU patients with aplastic anemia using machine learning techniques and constructed interactive nomograms to facilitate clinical decision-making. Using the MIMIC-IV database, we identified key clinical variables through a rigorous feature selection process, reducing more than 400 potential predictors to seven essential characteristics. The logistic regression model demonstrated superior predictive performance compared to Cox regression, achieving higher AUROC values across 7-day, 14-day, and 28-day mortality predictions.

To assess the generalizability of the model, an external validation was performed using the eICU database, where the performance remained robust despite a slight decrease in AUROC values (approximately 10%). This suggests that our model retains predictive capability in different cohorts of patients and clinical settings, reinforcing its potential for broader clinical application.

SHAP analysis provided insight into the relative contributions of key predictors, highlighting APS III, bicarbonate, and platelet count as crucial determinants of short-term survival. The importance of features varied over timeframes, with metabolic and hematologic parameters playing a more significant role in the prediction of early mortality, while systemic severity became more influential over longer durations.

The importance of permutation quantifies the contribution of each characteristic by measuring the decline of AUROC after swathering its values. APS III emerged as the strongest predictor in all models, underscoring its role in the evaluation of the severity of the ICU.

An interactive nomogram was developed to provide real-time, individualized risk assessments for ICU patients with aplastic anemia. This tool offers a user-friendly interface for clinicians, enabling rapid mortality risk estimation and improving critical care resource allocation.

In general, this study demonstrates the feasibility of applying machine learning and nomogram-based approaches to improve the prognostic assessment in patients in the ICU for aplastic anemia. Future work should explore dynamic modeling strategies, incorporating real-time physiological data and multicenter data sets to further enhance predictive accuracy and clinical applicability.

## Data Availability

All data produced in the present study are available upon reasonable request to the authors

## Acknowledgments

The authors express their appreciation to the developers of MIMIC-IV and e-ICU for providing a comprehensive and detailed public EHR dataset.

## Notes

### Competing Interest Statement

The authors have declared no competing interest.

### Funding Statement

This study did not receive any funding

### Author Declarations

The study used ONLY openly available human data from the MIMIC-IV and eICU databases, which were publicly accessible before the initiation of this study. These datasets are maintained by the MIT Laboratory for Computational Physiology and require registration and approval for access. MIMIC-IV Database: Available at https://physionet.org/content/mimiciv/2.2/ eICU Collaborative Research Database: Available at https://physionet.org/content/eicu-crd/2.0/ Access to these datasets requires completing the necessary training and obtaining approval via the PhysioNet Credentialing Process, ensuring compliance with data use agreements and ethical guidelines.

